# Mathematical modeling of the COVID-19 prevalence in Saudi Arabia

**DOI:** 10.1101/2020.06.25.20138602

**Authors:** Tusneem Elhassan, Ameera Gaafar

## Abstract

The swift precautionary and preventive measures and regulations that were adopted by the Saudi authority has ameliorated the exponential escalation of the SARS-CoV-2 virus spread, decreased the fatality rate and critical cases of COVID-19. Understanding the trend of COVID-19 is crucial to establishing the appropriate precautionary measures to mitigate the epidemic spread. The aim of this paper was to modifying and enhancing the mathematical modeling to guide health authority and assist in an early assessment of the epidemic outbreak and can be utilised to monitor non-pharmaceutical interventions (NPIs). Both ARIMA model and Logistic growth model were developed to study the trend and to provide short and long-term forecasting of the prevalence of COVID-19 cases and dynamics. The data analyzed in this study covered the period between 2^nd^ March and 21^st^ June 2020. Two different scenarios were developed to predict the epidemic fluctuating trends and dynamics. The first scenario covered the period between 2^nd^ March and 28^th^ May when the first peak was observed and immediately declined. The analysis projected that the COVID-19 epidemic to reach a peak by 17^th^ May with a total number of 58,534 infected cases and to end on the 4^th^ August, if lockdown were not interrupted and folks followed the recommended personal and social safety guidelines. The second scenario was simulated because of the sudden sharp spike witnessed in the trend of the new confirmed cases on the last week of May and continue to escalate till the time of current writing-21^st^ June. In the 2nd scenario, the analysis estimated the epidemic to peak on 15^th^ June with a total number of 146,004 infected cases and to end on 29^th^ September, 2020 with a final size of 209,607 (185,757 to 244,310) infected cases, assuming that the NPIs will be maintained while new normal life is resumed carefully. ARIMA and Logistic growth models showed excellent performance in projecting the epidemic prevalence, trends and dynamics at different phases. In conclusion, the analysis presented in this paper will assist policy-makers and health care authorities to evaluate the effect of the NPIs applied and to size the resources needed to manage different phases and cope with the final size of the epidemic estimates and to impose extra precautions.

## 1. Introduction

A global health crisis is resulting from the pandemic of the novel Coronavirus disease 2019 (COVID-19) that is caused by the severe acute respiratory syndrome coronavirus-2 (SARS-CoV-2) and originated from Wuhan, China. The WHO declared on January 30 as PHEIC and a pandemic on March 11, 2020 (1). As of, June 21, 2020, Covid-19 has spread to 213 countries with over 8 million confirmed cases and death claimed thousands of lives where 161,005 were reported in Saudi Arabia with more than 1,307 death. Decades of extensive experiences in management of Hajj and Umrah, have equipped the Kingdom with accumulated knowledge and expertise to deal with infectious epidemics promptly, as stated by Saudi health authorities (2). Additionally, dealing with MERS-CoV for the last seven years, resulting in a noticeable improvement in infection control practice in health care institutions in the Kingdom (3, 4). The government of Saudi Arabia implemented fast and stringent non-pharmaceutical interventions (NPIs) to prevent the spread of SARS-CoV-2 via human-to-human transmission (5). The government started with implementing a partial lockdown, and subsequently a full lockdown was implemented which led to a significant decline in the numbers of infected and mortality cases per day in Saudi Arabia (3, 6). Currently, there is no effective treatment to COVID-19 so, eradication of the epidemic and safe return to a healthy normal life will happen by the introduction of a vaccine or development of herd immunity which might take several months. Lockdown cannot be maintained for a long time as it creates significant societal and economic disruption. Thus, the focus of this research was to utilise ARIMA model and Logistic Growth model to provide short and long-term forecasting and to estimate the COVID-19 dynamics in order to assist the health authority on their strategic planning and resource allocation of COVID-19 in Saudi Arabia. Mathematical models can provide guidance and help in the early assessment of the epidemics outbreak and evaluate the NPIs (7, 8). Additionally, they can facilitate the initial estimation of the final scale of the epidemic to assess needs for treatment and to compare the effects of different treatment approaches (9). It is essential to be able to project the final extent of the epidemic to assess the healthcare resources and interventions needed to efficiently cope with the outbreak in different phases. Comprehending the prevalent trend of COVID-19 is crucial to establishing the appropriate precautions to control the epidemic spread. Different mathematical models were used to estimate the COVID-19 dynamics in the literature depending on the stage of the epidemics. Exponential Logistic growth model was utilized to study the pattern of COVID-19 epidemics in early stages in KSA (6). It has also been utilized to model late COVID-19 stages in China (10). Some mathematical models are appropriate for early stages while others fit data associated with later stages of the outbreak. For example, both exponential and logistic models have been applied to the early stages. However, exponential models show unreasonable predictions at late stages. Alternatively, logistic models are more suitable for modelling late stages of the epidemic as new cases start to decelerate when approaching the maximum capacity limit (11, 12). Komies et al.(6), utilized the Exponential Logistic Growth and Susceptible Infectious Recovered (SIR) models to estimate and compare the COVID-19 dynamics in KSA and UK in the very early phase of the epidemic before the introduction of active screening. The epidemic final size and growth rate were estimated to be only 2064 and 0.2, respectively, utilizing data until the first of April. Ketcheso et al. (13), developed an SIR model to estimate past, current, and future COVID-19 infections using recorded COVID-19 deaths on age-specific population in KSA and other counties. Alshammari et al. (14), studied the effect of interaction with infected people (symptomatic & asymptomatic) on the COVID-19 pandemic in KSA and estimated the maximum number of hospital and ICU beds required during peak time. The governing mathematical models such as Autoregressive Integrated Moving Average (ARIMA) have been utilised for short-term forecasting of COVID-19 cases with excellent results. Alzahrani et al. (15) developed an ARIMA model to forecast COVID-19 confirmed cases in Saudi Arabia from April 21^st^ to May 21^st^. However, recovery and death were not included in his projections as he used data from the early phase of the outbreak till the April 21^st^. Furthermore, ARIMA models have been utilised to display the trends of COVID-19’s prevalence in Italy, Spain and France (16). Chintalapud et al. (17) applied AIRMA model to forecast the confirmed infected and recovered COVID-19 cases in Italy after sixty days of lockdown. Bayyurt et al.(18) has applied AIRIMA model to forecast COVID-19 cases and deaths in Italy, Spain and Turkey. Tandon et a (19) has developed an ARIMA model to forecast the COVID-19 cases in India. However, others implemented logistic growth model to estimate COVID-19 dynamics. Shen CY (10) utilised Logistic Growth model to predict the new COVID-19 cases using non-linear least square method in multiple regions in China and nations with a large amount of cases. A generalized logistic growth model was applied to estimate the trend of the dynamics of COVID-19 in 29 provinces of china, four severely affected countries (Iran, South Korea, Italy and Japan), and Europe as a whole (12). In this study, the prevalence, and dynamics of COVID-19 outbreak in Saudi Arabia covering the period from 2^nd^ March to 21^st^ June, were estimated using ARIMA and Logistic Growth models. ARIMA models were utilised to study the trend and to provide short-term forecasting of the prevalence of COVID-19 confirmed, recovered and death cases in KSA. While, logistic growth model was used to estimate the COVID-19 dynamics such as the final size, maximum growth rate, peak and expected end-time. In order, to deal with the trend oscillation of COVID-19 cases in the last three months, models were developed to predict the epidemic dynamics for two scenarios embracing different phases. The first scenario covered the period between 2^nd^ March and 28^th^ May when the first peak was observed and incidence of new cases started to decline, and the model predicted the epidemic to end on August 4, 2020 if the lockdown and curfew were not interrupted. The second scenario covered the period between 29^th^ May and 21^st^ June where a sudden jump in COVID-19 new cases was witnessed due to lessening of the lockdown and non-compliance of folks to safety rules and authority guidelines upon gatherings. Analysis of the 2^nd^ scenario projected the pandemic to end on September 29, 2020. Consequently, our model provided estimates for dynamics if the interventions were completely relaxed. This research has two complementary objectives: (1) to provide short-term forecasting of the prevalence of COVID-19 confirmed, recovered and deceased cases. (2) to estimate the dynamics in terms of the epidemic growth rate, final size, peak time and ending time. Hopefully, the analysis and the forecasts provided in this manuscript will assist the health authority, in their preparation and strategic planning to combat COVID-19 spread and allocate the resources needed at peak times.

## Models and Methods

The prevalence and incidence of COVID-19 was taken from the World Ministry of Health (WHO), Saudi Ministry of Health (MOH), other MOH websites such as Sehhtty and the Saudi CDC (1, 20-22).This paper investigates two models and analyze the results numerically using some indicators of performance evaluation.

### 1.1 ARIMA model

ARIMA model is a statistical model that uses time series data to study the trend and generate future forecasting of a time series data. A time series is a sequence of time-ordered numerical data-points taken in equally spaced points of time (23). Time series forecasting is the use of modeling to predict future values based on previously observed values (24). ARIMA models are well suited to short-term forecasting with a variety of time series. They take into consideration several essential features of time series such as the trend and periodic changes as well as random disturbance (16). ARIMA models was first introduced by Box and Jenkins in 1970 and became one of the most applied models in time series analysis. The model is expressed as an ARIMA(p, d, q) where p is the order of autoregression, d is the degree of difference, and q is the order of moving average (24). ARIMA model is a generalized model that integrates the autoregressive model AR(p) and the moving average model MA(q). It can also be reduced into other simpler models such as ARMA (p,q), AR(p) and MA(q) models. ARMA is a stationary model which does not require differencing. Sequence differencing is required to achieve stationarity. ARIMA models that do not require differencing are considered as ARMA models. AR (p) is an autoregression model of order p which means that the current time series values linearly depend on the earlier *p* values of the time series and the current residual. On the other hand, MA (q) is a linear regression model of the current value of the time-series against current and earlier residuals known as white noise.

The AR(p), AR (q) and ARMA models can be expressed as polynomials of autoregressive, residuals and a combination of them Equations: (1), (2) and (3) respectively.

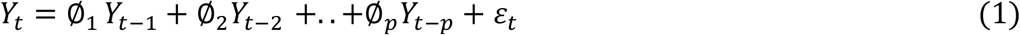

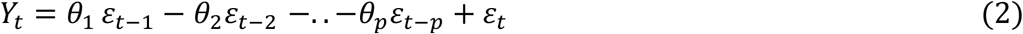

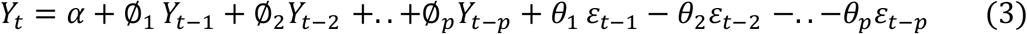

Where ϕ and θ are the autoregressive and moving average parameters, respectively. *Y*_*t*_ is the observed time series value at time t, *ε*_*t*_ = *Y*_*t*_ − *Y*_*t*−1_, is the random shock at time t which assumed to be normally distributed with a mean of zero and variance of *σ*^2^ and α is a constant. ARIMA and ARMA models are represented using the same equation, equation (3), (16).

#### Model evaluation

Models were compared based on their goodness of fit using Akaike information criteria. The Model with the minimum AIC value was selected. AIC was used to calculate the amount of information loss depicted by the fitted model, and can be defined as follows:

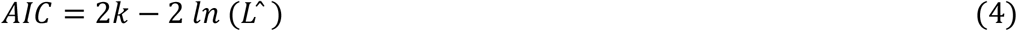

Where k is the number of model parameters and 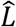 is the maximum model likelihood value estimated by the maximum likelihood function, and n is number of observations.

Model performance was evaluated, and the best model was selected based on four measurements, namely Root Mean Square Error (RMSE), Mean Absolute Error (MAE), Mean Absolute Percentage Error (MAPE) and coefficient of determination (R^2^). Equations (5), (6) and (7) were used to calculate the RMSE, MAE and MAPE respectively (10).

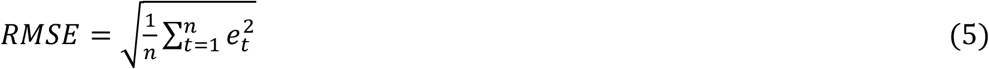

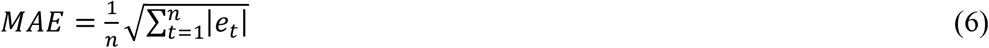

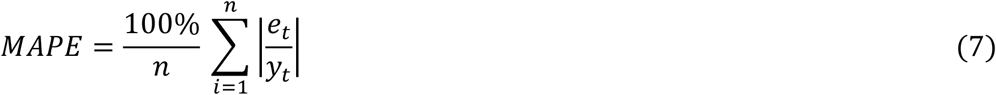

where y_t_ is the observed value at time t, e_t_ is the difference between the observed and predicted values at time t. Also, n is a sequence of time points. Lower RMSE, MAE, and MAPE values indicate a better calibration and therefore better performance. Analyses were performed using was performed using R studio version 1.2.5033 and IBM SPSS statistics version 24. P-value <0.05 was considered significant.

### 1.2 Logistic growth model

Logistic function has several applications in a variety of fields including biology, economics, and epidemiology (25). In the field of epidemiology, logistic growth function has been applied in molding the population growth and epidemic outbreak. The logistic growth is characterized by an increasing growth rate at the beginning of epidemic, however, as the population sized increases and approaches the maximum limit, the growth rate starts to decrease until it reaches zero and the curve change from a convex to a concave. The maximum limit is called the carrying capacity and it is defined as the maximum number of cases that can be supported by the environment according to the limited resources (26). The logistic regression function takes the following form:

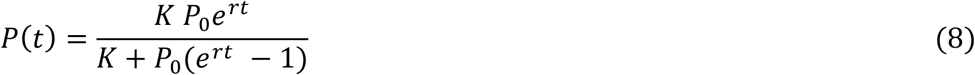

Where P(t) is the cumulative number of COVID-19 cases, K is the carrying capacity, t is epidemic time frame, r is the COVID-19 growth rate and P_0_ is the initial value of function at time (t_0_= 0).

In this study, the logistic growth model derivative has been used to model the new COVID-19 cases using the following equation:

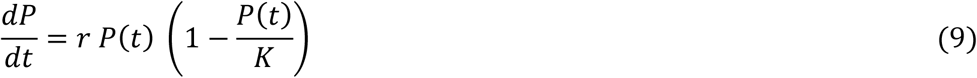

By substituting (8) in (9), equation (9) can be written as follows:

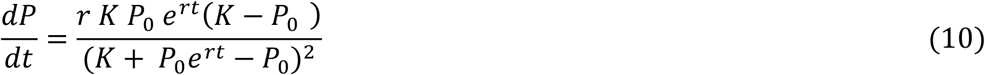

Equation (10) is the logistic growth differential equation expressed as a function of r, K and P_0_ f (r, K, P_0_) where 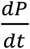 represents the number of new COVID-19 cases. Therefore, equation (10) can be written as the following:

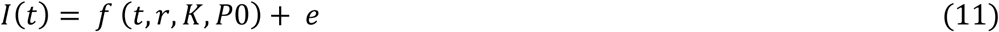

Parameters *r, K, P*_0_ can be estimated by minimizing the following sum of square errors as follows:

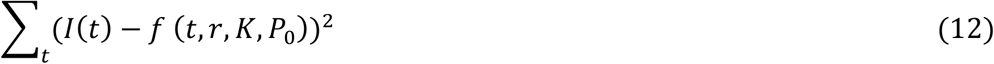

Equation (12) is a minimization problem that was solved using Non-Linear Square method (NLS)(10).

The peak time (*t*_*peak*_), ending (*t*_*end*_), total number of infected cases at peak time, and new incidence at peak time (*dC*_*peak*_) of COVID-19 were estimated using the following equations(27).

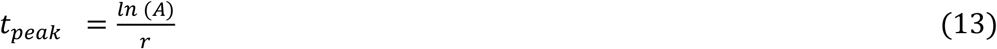

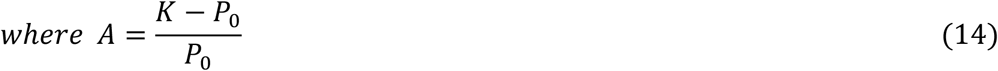

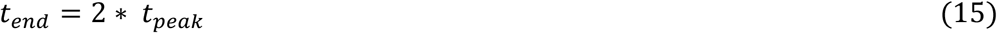

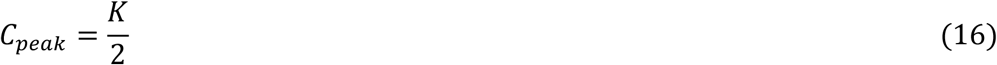

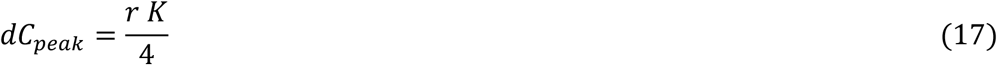

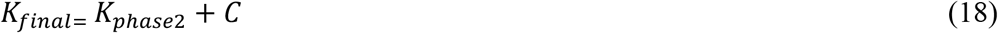

In equation (19), *K*_*final*_ is the COVID-19 final size, *K*_*phase*2_ is phase 2 final, and C is a constant representing the total number of infected people at the beginning of phase 2 of COVID-19 in SA. However, the minimum level of *K*_*final*_ is 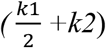, while the maximum value is (k1+ k2).

#### Model evaluation

The model accuracy was evaluated using the coefficient of determination (R^2^) calculated as follows:

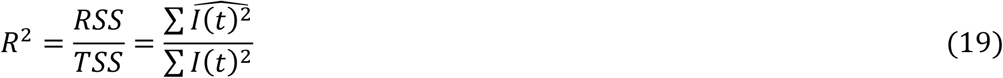

where RSS represents the Residual Sum of squares, 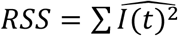, and TSS presents the Total Sum of squares, *TSS* = ∑ *I*(*t*)^2^.

### 2. Implementation

Models were implemented using 64-bit operating system, x64-based processor, 16.GB RAM, and Intel® Core ™ i7-9750h @2.60GHz., windows 10 home operating system and R studio version 1.2.5033 and IBM SPSS statistics version 24. Fig. (1a &1b) shows a flow chart for implementation Logistic Growth and ARIMA models.

**Fig.1a& 1b:**
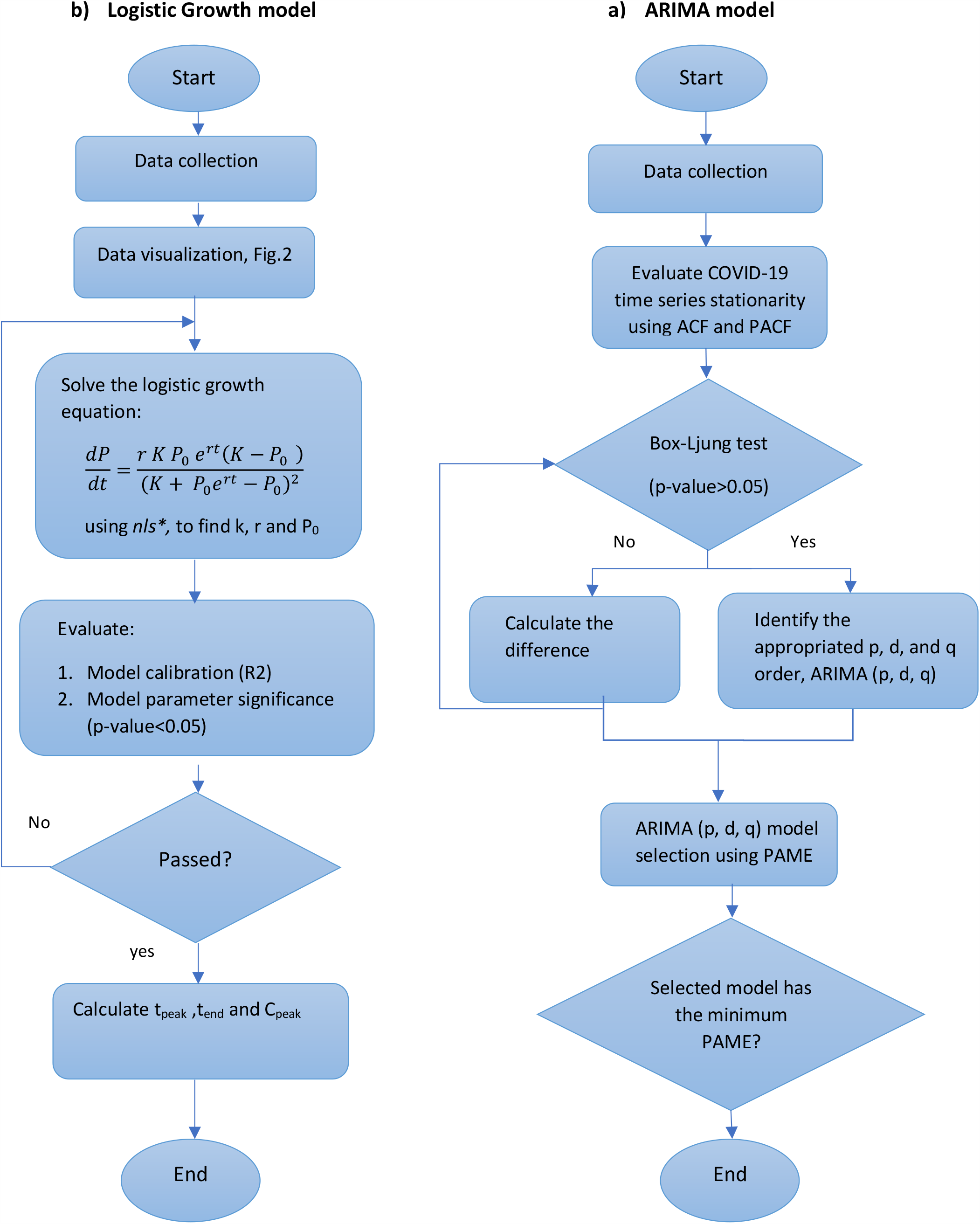
Represents the logistic growth model and the ARIMA model, respectively. *nls\** is the non-linear least squares function.

## 3. Results and discussion

### 3.1 Descriptive statistics of COVID-19 time series

Descriptive statistics of the prevalence of confirmed COVID-19 cases between 2^nd^ March 2020 and 21^st^ June 2020 are given in Table 1. The prevalence of COVID-19 confirmed, death, and recovered cases reported during this period were 161,005, 1307, and 105175, respectively.

**Table 1:** descriptive statistics of COVID-19 incidence and prevalence

|  | N<br>Statistic | Minimum | Maximum | Mean | Std.<br>Deviation | Skewness |  | Kurtosis |  |
| --- | --- | --- | --- | --- | --- | --- | --- | --- | --- |
|  |  | Statistic | Statistic | Statistic | Statistic | Statistic | Std.<br>Error | Statistic | Std.<br>Error |
| * new cases | 112 | 0 | 4919 | 1437.54 | 1341.9 | 0.72 | 0.228 | -0.376 | 0.453 |
| **confirmed | 112 | 1 | 161,005 | 40898.8 | 47261.5 | 0.99 | 0.228 | -0.203 | 0.453 |
| **Death | 112 | 0 | 1,307 | 282.9 | 345.5 | 1.4 | 0.228 | 1.113 | 0.453 |
| **Recovery | 112 | 0 | 105,175 | 23898.4 | 32923.1 | 1.1 | 0.228 | -0.316 | 0.453 |

\*Incidence. \*\*Prevalence of confirmed, recovered and dead cases.

### 3.2 ARIMA Model

#### Development of ARIMA model

Development of ARIMA model involves four iterative phases namely, 1) assessment, 2) parameter estimation, 3) model diagnostics, and 4) forecasting.

#### Model assessment

Stationarity and seasonality of COVID-19 time series were first assessed using autocorrelation function **(ACF)** and partial autocorrelation function **(PACF)** correlogram’s. Stationarity is an important assumption to obtain accurate future predictions. A time series is considered stationary if its statistical properties such as mean, variance and autocorrelation structure are constant over time (28). The trend lines of COVID-19 time series incidences and prevalence’s during the time between 2^nd^ March to 21^th^ June 2020 are shown in Fig. (1a& 1b), respectively. ACF calculates the autocorrelation of the current time series observation with its lagged values. While, PACF calculates the correlation of the residuals after removing the effects explained by the earlier lags. ACF and PACF correlogram’s are used to identify the numbers of AR and MA terms that are needed in the ARIMA model. ACF and PACF correlograms of the COVID-19 series total confirmed, death and recovered cases are showed on Fig. (3a &3b, 3c &3 d, 3e & 3f), respectively.

#### The trend of COVID-19 series

By looking at Fig. 1a, the COVID-19 epidemic was slowly growing and then started to accelerate with time. It reached a first peak on 15^th^ May 2020 then declined, however, since 29^th^ May 2020 the COVID-19 cases started to exponentially accelerate again, and a spike was noted. ACF shows a significant autocorrelation of the COVID-19 time series prevalence, Fig (2a). The straight lines in Fig. (3a & 3b) represent the 95% confidence interval limits of the non-zero autocorrelation. The bar boxes are extended beyond the confidence interval lines indicating a significant autocorrelation and evidence of non-stationarity. Box-Ljung test shows a significance autocorrelation between the COVID-19 time series observations and their earlier lags (p-value<0.05) therefore, first-order difference was calculated. The second-order difference was furtherly calculated as the time series was still showing significant autocorrelation. However, after applying the second-order difference, all the time series were found to be stationary and no significant autocorrelation was observed (p-value >0.05), Fig. (S1.a, S1.b & S1.c). COVID-19 has also showed no evidence of seasonality, Fig. (2a& 2b, 3a & 3b).

**Fig. 2a & 2b.**
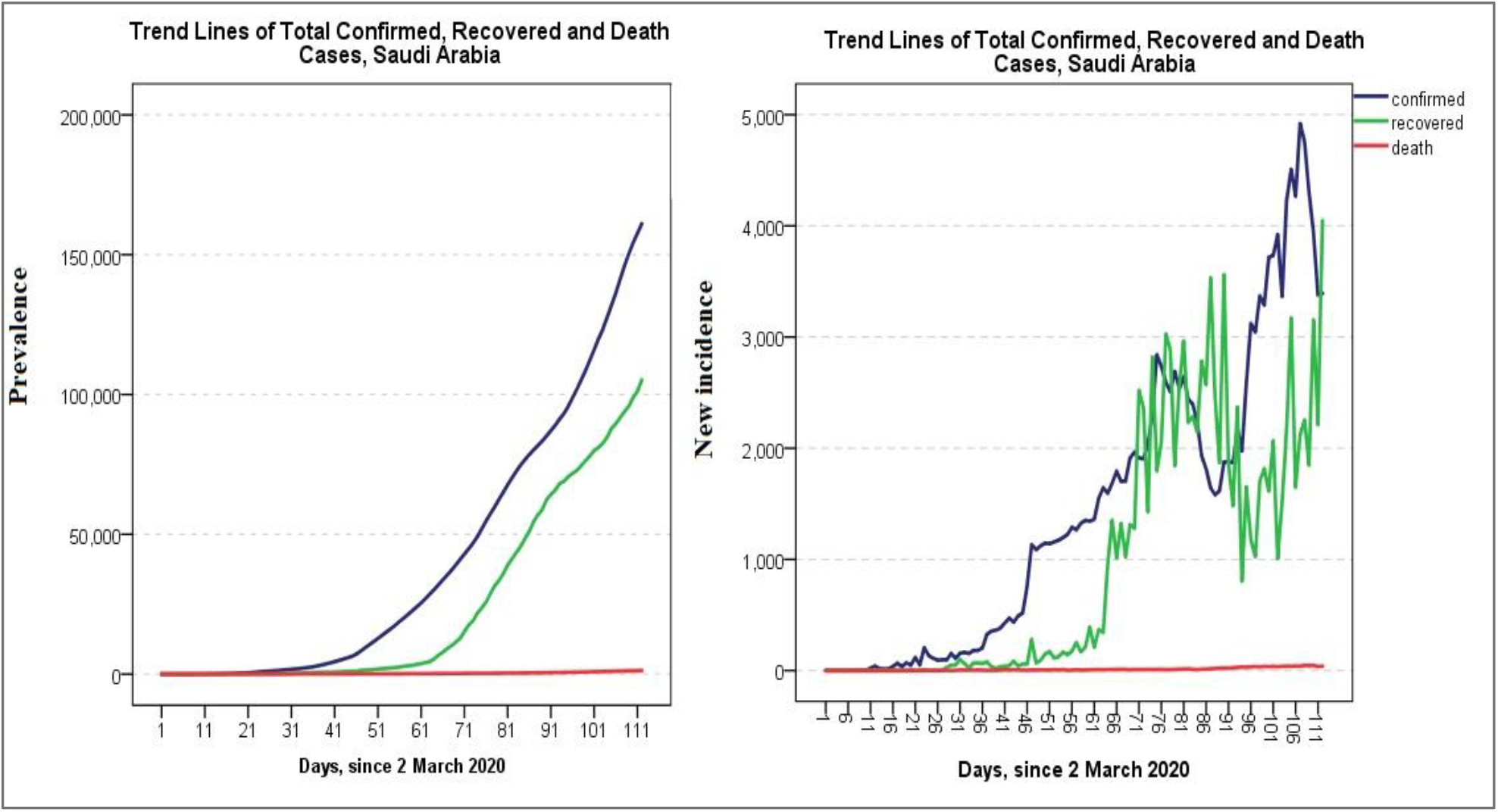
Shows the incidence and prevalence of the COVID-19 confirmed, recovered, and death cases between 2^nd^ of March and 21^st^ of June 2020 in the Kingdom of Saudi Arabia.

**Fig. (3a &3b, 3c &3d, 3e & 3f):**
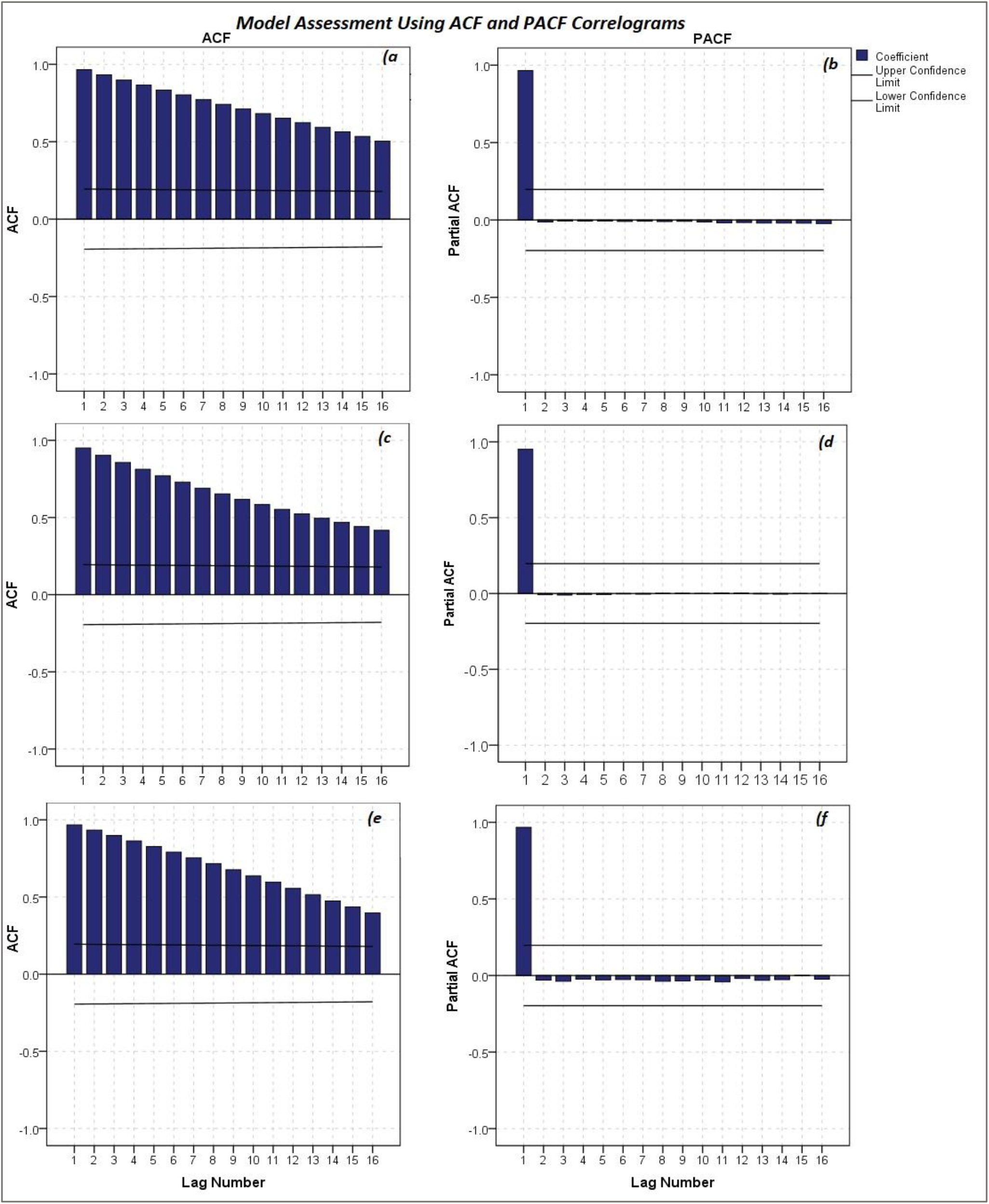
Shows ACF and PACF correlograms of the COVID-19 total confirmed, death and recovered cases, respectively.

#### Model fitting and parameter estimation

Model fitting involves three compartments, namely prevalence of confirmed cases, death, and recovery.

##### 1. Prevalence of COVID-19 confirmed cases

ARIMA (0,2,0) was selected to fit the prevalence of COVID-19, which is mostly used to fit time series that has a changing trend over time (Fig.2a). In this study, ARIMA (0,2,0) was used to adjust for the changing trend of the COVID-19 time series in Saudi Arabia. In this model, a new observation can be predicted based on the current observation and the most recent change of the COVID-19 trend. The model performance was evaluated based on the “goodness of fit” using AIC. Furthermore, statistical measures such as (RMSE), (MAE) (MAPE) were also utilised and the model with the minimum AIC and minimum MPAE was selected, Fig.S2. ARIMA (0,2,0) showed an AIC of 1484.6, MAPE of 4.2, and 95.7% accuracy. A 10-days forecasting of COVID-19 confirmed cases is shown in table (2 &3), Fig.4.

**Fig. 4.**
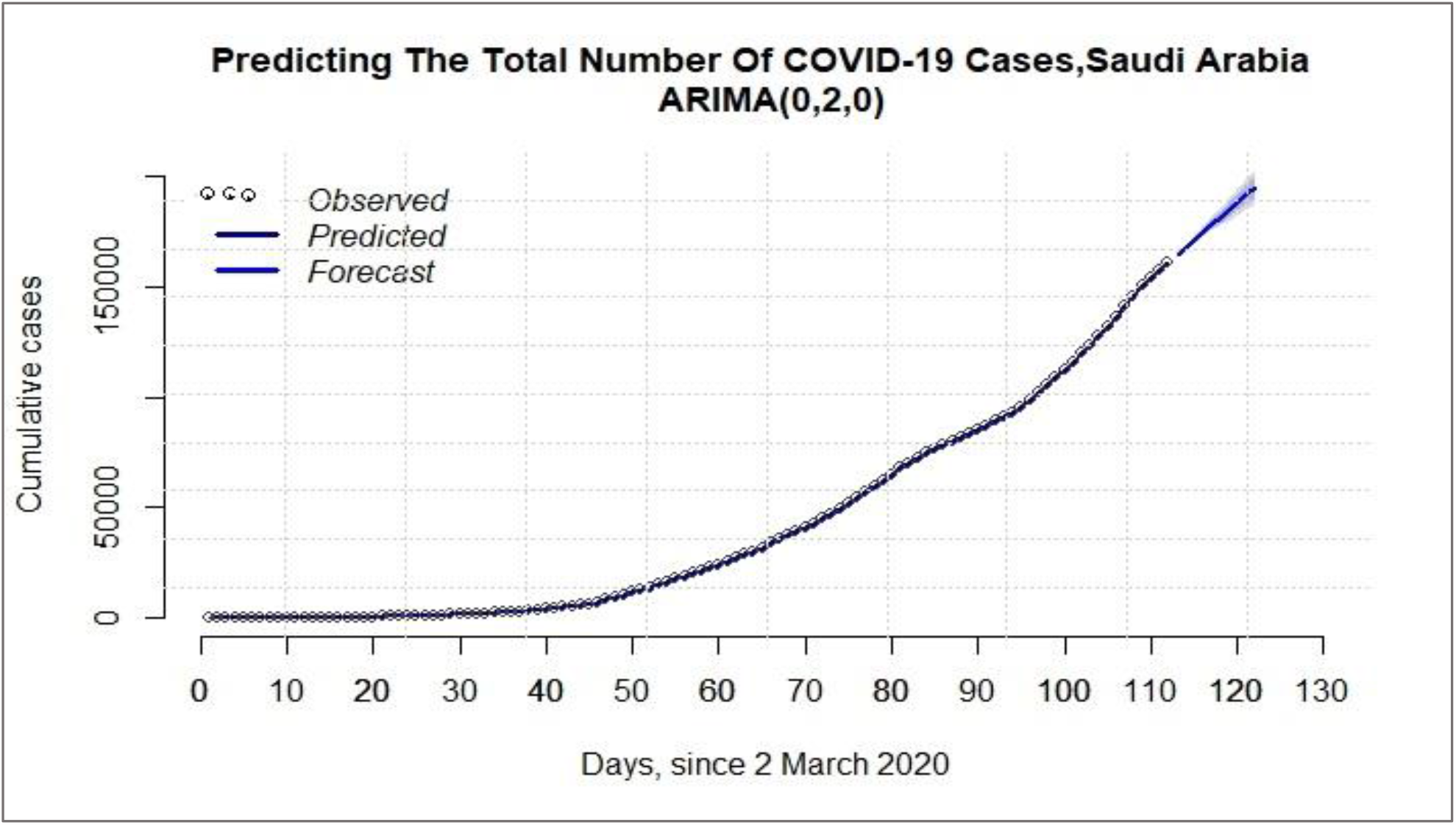
Prediction of the prevelence of COVID-19 confirmed cases in Saudi Arabia by ARIMA (0,2,0) model. The solid dark blue line represents the predicted data and the open circles represents the obseverd data. The light blue line presents the the prevelance forcast up to 1^st^ of July and the shaded area shows the the 95% confidence interval.

**Table 2:**
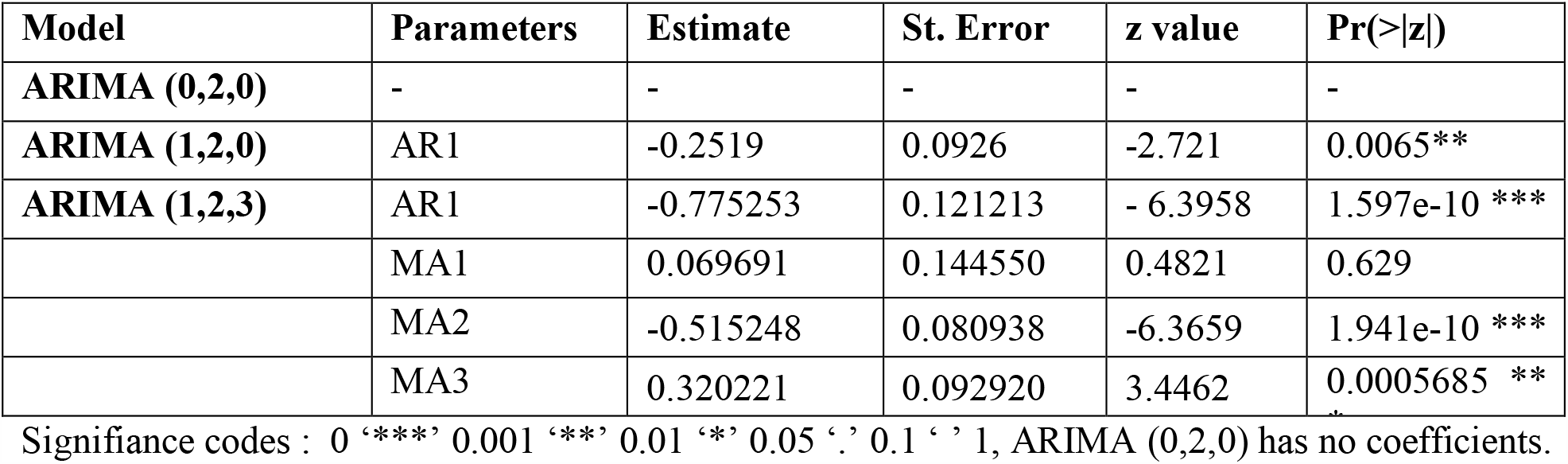
ARIMA (p,d,q) model parameters

**Table 3:**
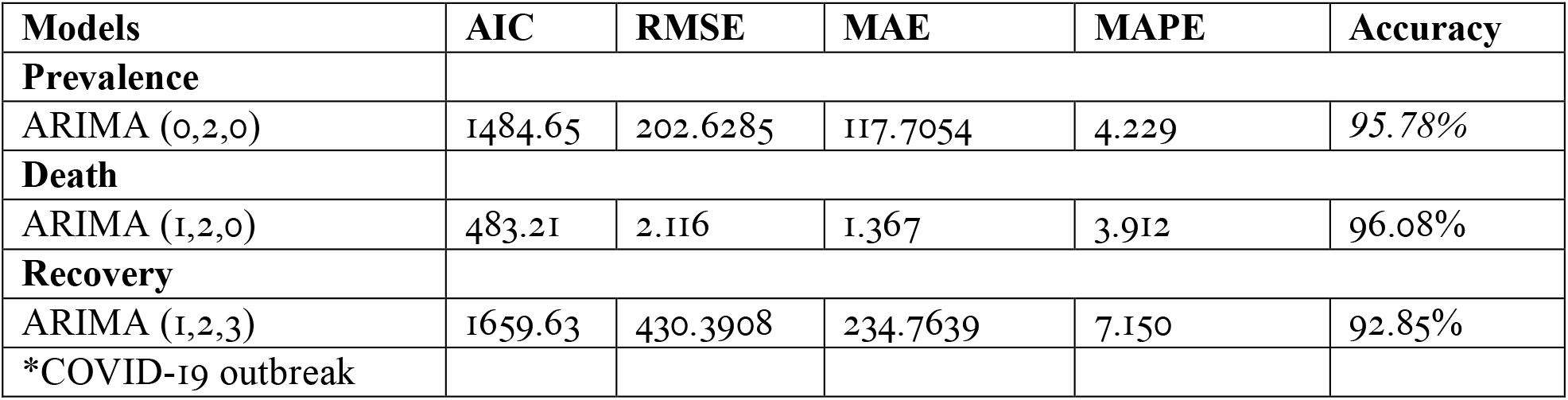
ARIMA (p,d,q) models for perveance of COVID-19 cases, death, and recovery

##### 2. Prevalence of COVID-19 death

ARIMA (1,2,0) was also selected to model the prevalence of COVID-19 death indicating a significant association with its first lagged value, Fig.S3. The model showed a good calibration, AIC=483.2 and PMAE= 3.9, table (2&3), Fig.5.

**Fig. 5.**
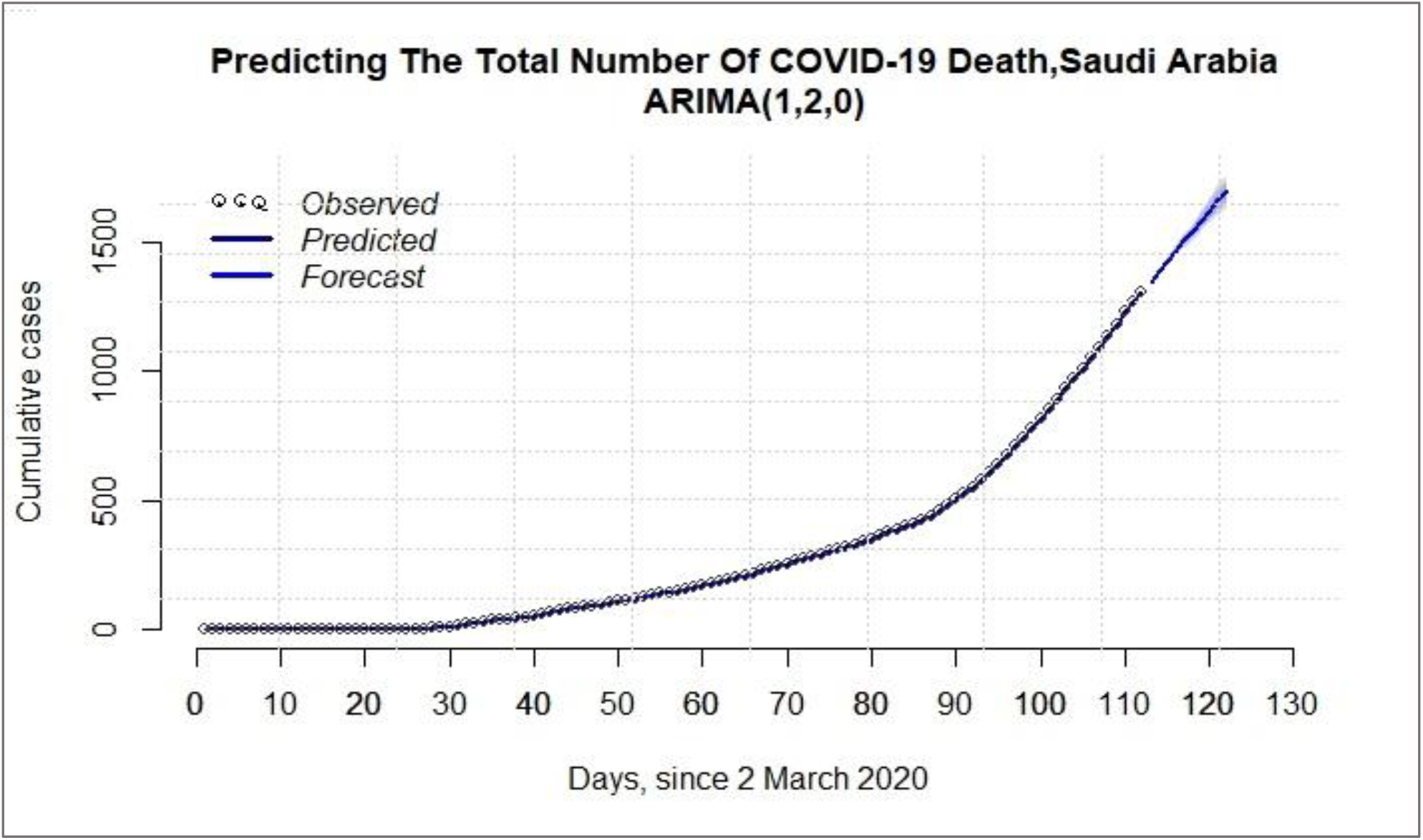
Prediction of the prevelence of COVID-19 death in Saudi Arabia by ARIMA (1,2,0) model. The solid dark blue line represents the predicted data and the open circles represents the obseverd data. The light blue line presents the the death forcast up to 1^st^ of July and the shaded area shows the the 95% confidence interval.

##### 3. Prevalence of COVID-19 recovered cases

ARIMA (1,2,3) was selected to model the prevalence of COVID-19 recovery which indicates that the prevalence of COVID-19 cases was significantly associated with its first lagged value, and weighted moving average of the past three forecast errors, Fig.S4. The model showed a good calibration, AIC=1659.63 and MPAE= 7.15, table (2&3), Fig.6.

**Fig. 6.**
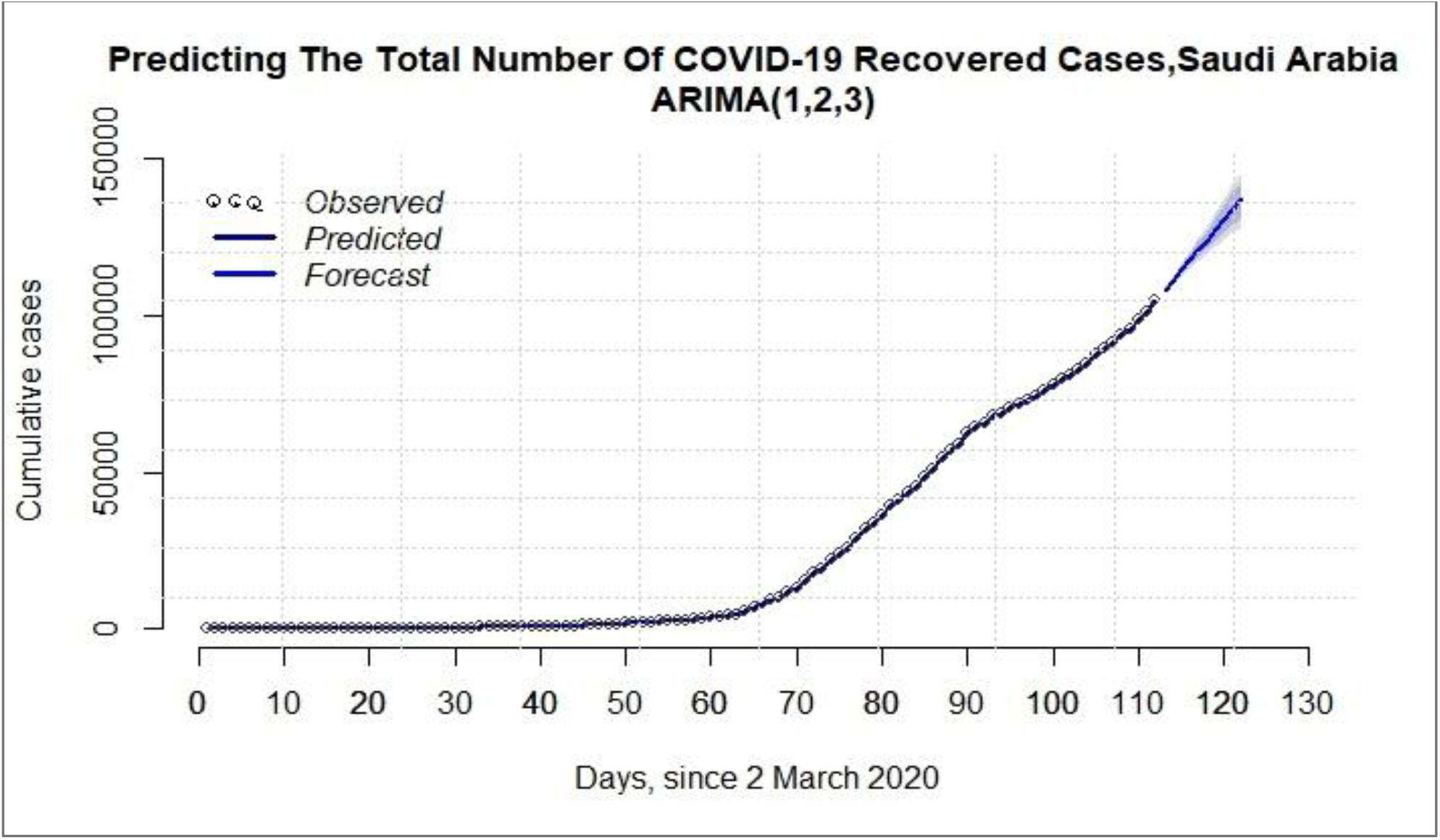
Prediction of the prevelence of COVID-19 recoved in Saudi Arabia by ARIMA (1,2,3) model. The solid dark blue line represents the predicted data and the open circles represents the obseverd data. The light blue line presents the the recovered forcast up to the 1^st^ of July and the shaded area shows the the 95% confidence interval.

#### Forecasting

The forecast of the ARIMA models for the successive 10-days of total cases is presented in table (4). The forecast indicated gradual increase in the reaching 194,935; 1,700; and 137,224 for total confirmed cases, death, and recovery by 1^st^ July 2020, respectively. Up to 1^st^ July, the forecast estimated the percentage of the total recovered, and death case to be 70.3% and 0.87% respectively.

**Table 4:** 10-days forecast of new COVID-19 cases in Saudi Arabia

| Date | Day | Forecast | 95%. | 95%. | Death | 95%. | 95%. | Recovery | 95%. | 95%. |
| --- | --- | --- | --- | --- | --- | --- | --- | --- | --- | --- |
| 22-Jun-20 | 113 | 164398 | 163997.3 | 164798.7 | 1346 | 1342.0 | 1350.4 | 108046.1 | 107179 | 108913.2 |
| 23-Jun-20 | 114 | 167791 | 166894.9 | 168687.1 | 1385 | 1377.2 | 1394.1 | 111129.5 | 109711.2 | 112547.8 |
| 24-Jun-20 | 115 | 171184 | 169684.6 | 172683.4 | 1425 | 1411.3 | 1438.76 | 114515.3 | 112518.7 | 116511.9 |
| 25-Jun-20 | 116 | 174577 | 172382.1 | 176771.9 | 1464 | 1444.7 | 1484.1 | 117666.7 | 114879.8 | 120453.6 |
| 26-Jun-20 | 117 | 177970 | 174998 | 180942 | 1503 | 1477.5 | 1530.1 | 120999.8 | 117396 | 124603.6 |
| 27-Jun-20 | 118 | 181363 | 177540.2 | 185185.8 | 1543 | 1509.6 | 1576.8 | 124192 | 119641 | 128743.1 |
| 28-Jun-20 | 119 | 184756 | 180014.4 | 189497.6 | 1582 | 1541.2 | 1624 | 127493.5 | 121956.2 | 133030.8 |
| 29-Jun-20 | 120 | 188149 | 182425.3 | 193872.7 | 1622 | 1572.3 | 1671.7 | 130710.3 | 124093.5 | 137327 |
| 30-Jun-20 | 121 | 191542 | 184776.8 | 198307.2 | 1661 | 1602.9 | 1719.9 | 133992.7 | 126251.6 | 141733.8 |
| 01-Jul-20 | 122 | 194935 | 187071.9 | 202798.1 | 1700 | 1633.0 | 1768.6 | 137224.2 | 128285.5 | 146162.9 |

### 5.2 Logistic growth model

In this analysis, we postulated two different scenarios based on the fluctuation of prevalence movement observed due to diversities in the precautionary mediations that have been implemented by the government in different sequential phases, which were also interrupted with episodes of relaxation. Therefore, the analysis portrayed two scenarios: The first scenario related to the period from 2 March to 28^th^ May and the second scenario covered the whole period from 29^th^ May to 21st June. Logistic growth model has been independently developed to each scenario separately, Fig.6.

#### Estimating COVID-19 dynamics

Based on our model, the COVID-19 dynamics [final size, growth rate, peak time, the new incidence and prevalence at peak time, and the epidemic ending time] were estimated.

##### Scenario 1

Using the historical data of the daily confirmed cases during the period between 2^nd^ March to 27^th^ May,2020, and depicted as scenario one, we estimated final size was 171,105 cases. This indicates that the maximum number of the total COVID-19 cases might reach 171,105 with a maximum growth rate of 0.079. Our forecast estimated the peak-time to be reached on 17^th^ May 2020 with a total number of 58,534 infected and 2334 new incidence cases, table (5). The epidemic was expected to end by 4^th^ August. Logistic growth model showed an excellent goodness of fit (R^2^ =0.982) which means that 98.1% of the variability of the total estimated COVID-19 cases is explained by the model, Fig.7.

**Fig.7.**
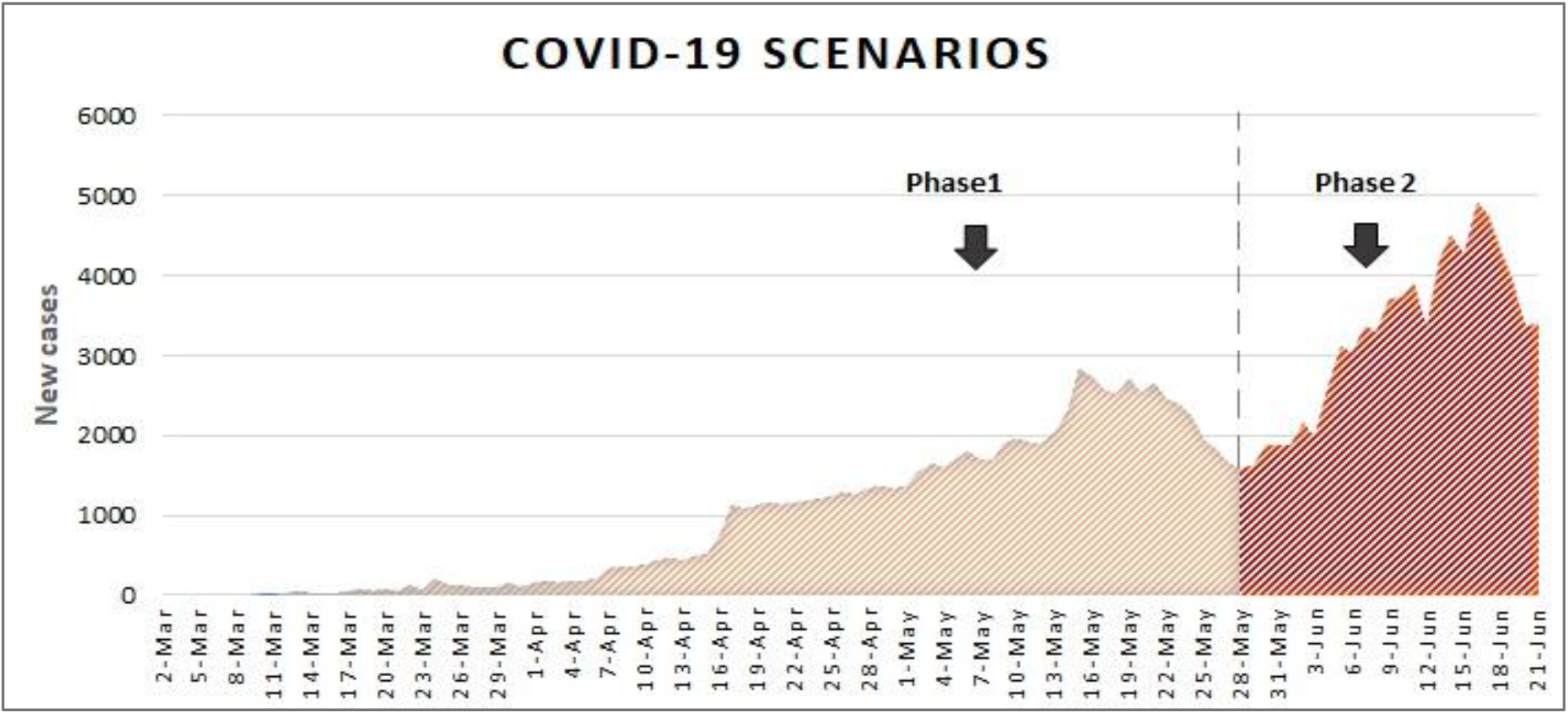
Shows the different phases of COVID-19 outbreak in Saudi Arabia. Phase 1 (2^nd^ March to 28^th^ May 2020) where partial to complete lockdown with other NPIs were implemented. While, phase 2 (28^th^ May to 21^st^ June) shows escalation of the COVID-19 confirmed cases coincide with the relaxation of lockdown and lessening of curfew were imposed.

**Table 5:** Parameters estimation of Logistic Growth Model (2 March 2020 to 27 May 2020)

| Parameters | Estimate | Std. Error | t value | Pr (>\|t\|) |
| --- | --- | --- | --- | --- |
| **r** | 7.977e-02 | 3.171e-03 | 25.16 | < 2e-16 *** |
| **K** | 1.171e+05 | 4.316e+03 | 27.12 | < 2e-16 *** |
| **P0** | 2.409e+02 | 5.696e+01 | 4.23 | 5.87e-05 *** |

Significance codes : 0 '\*\*\*' 0.001 '\*\*' 0.01 '\*' 0.05 '.' 0.1 ' ' 1

**Table 6:** Parameters estimation of Logistic Growth Model (2nd May to 11th June 2020)

| Parameters | Estimate | Std. Error | t value | Pr (>\|t\|) |
| --- | --- | --- | --- | --- |
| **r** | 1.354e-01 | 9.356e-03 | 14.47 | 2.14e-12*** |
| **K** | 1.272e+05 | 7.717e+03 | 16.48 | 1.72e-13 *** |
| **P<sub>0</sub>** | 7.477e-02 | 7.405e-02 | 1.01 | 0.324 |

Significance codes : 0 '\*\*\*' 0.001 '\*\*' 0.01 '\*' 0.05 '.' 0.1 ' ' 1

In table (5), k represents the maximum limit of COVID-19 total cases, r is the maximum per capita growth rate and P_0_ is the model initial value at time (t_0_=0).

##### Scenario 2

Scenario two cover the phase between 29^th^ May to 21^st^ June, 2020, the number of COVID-19 cases displayed a sudden exponential increase which might be attributed to the relaxation of curfews and lessening the lockdown that was implemented previously by the Saudi government. Accordingly, the final size of this phase was estimated to be 127,205 cases with a maximum growth rate of 0.13. Moreover, the peak-time is expected to be on 15^th^ June 2020 and the total number of infected people to be 63601. The incidence of new cases at peak-time was expected to be 4305 and the epidemic to end on September 29, 2020, table (5). Model showed good calibration, R2 =0.992, Fig.8. The total cumulative number of COVID-19 cases starting from 2^nd^ march till the 15^th^ June 2020 (peak 2) was expected to reach 146,004 and final size of the epidemic to be 209,607 (range: 185,757.5 -244,310).

**Fig. 8.**
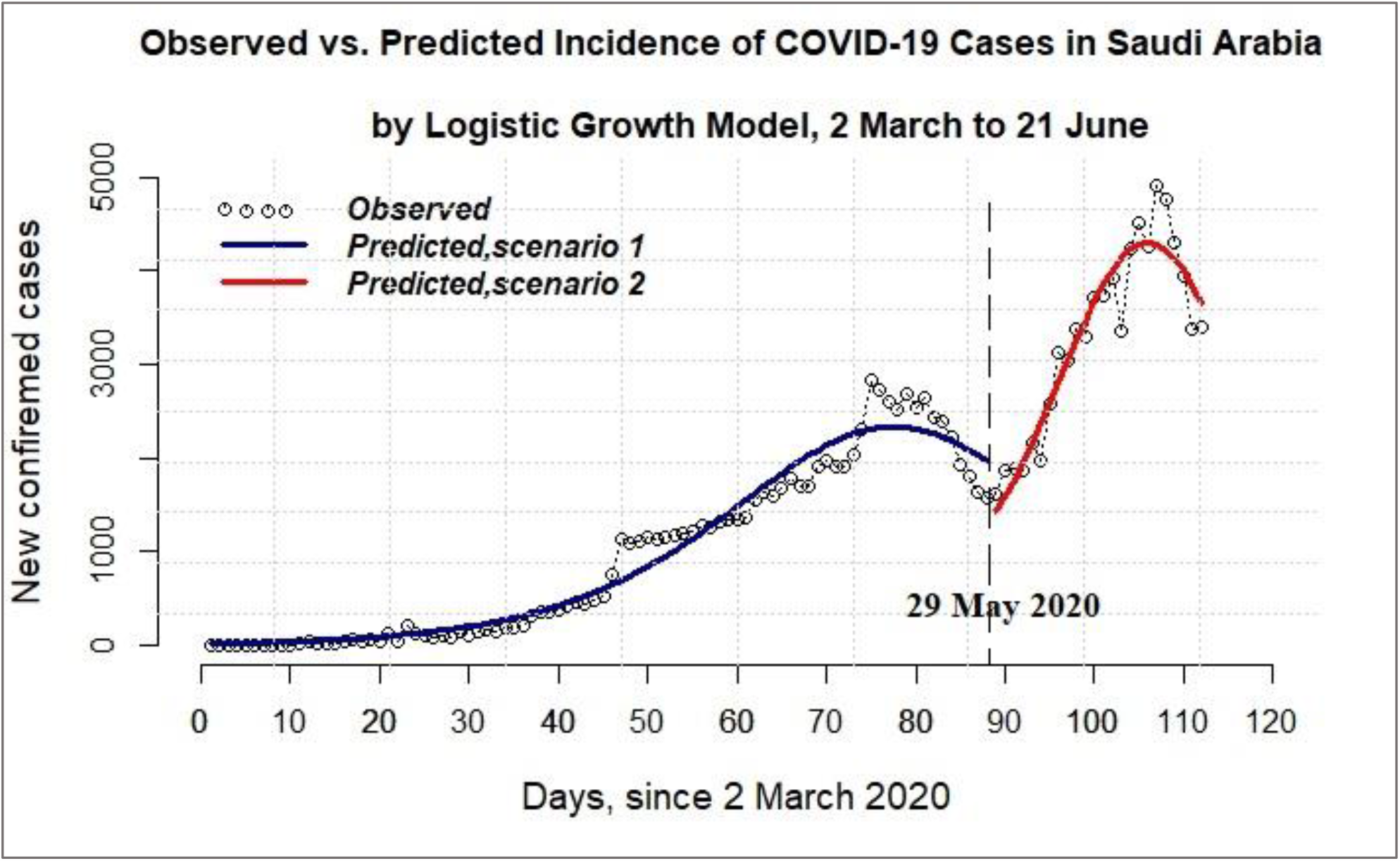
Illustrates the simulated scenarios covering phase1(2^nd^ March to 28^th^ May) and phase 2 (28^th^ March to 21^st^ June) in Saudi Arabia.

## Discussion

The pandemic of COVID-19 has an unfavorable effect on our daily life, health care system and the economics. The coronavirus (SARS-CoV) is a highly contagious disease that can spread rapidly and has claimed thousands of lives. The disease spread to the whole globe affecting more than eight million in 213 countries, and the death toll approaching half a million as of 21^st^ June 2020. It is terrifying to observe that the numbers of positive and fatality cases are increasing daily, which mandate groundbreaking means to be taken to win the battle over COVID-19,(21, 22, 29). At present, there is no effective cure or vaccine for COVID-19, and NPIs and mass surveillance to early discover positive cases are the only means to mitigate and suppress the virus spread. Therefore, understanding the dynamic and trend of COVID-19 is crucial to establish the appropriate precautionary measures to limit the epidemic consequences and to avoid overwhelming healthcare facilities, especially ICUs. The interventions implemented by the Saudi government in its battle against COVID-19 since 4^th^ March, included social engineered distance, isolation of infected cases, quarantine of their corresponding exposed cases (case-based) and non-case based (self-hygiene and lockdown),(5, 29). Dealing with MERS for the last seven years and running Hajj and Umrah for decades has equipped the Saudi health authority to respond effectively and rapidly to COVID-19. The swift precautionary measures and regulations that were adopted by the Saudi authority has ameliorated the exponential escalation of the virus spread, decreased the fatality rate and critical cases (30). Saudi health authority took fast actions and implemented strict precautions even before the report of the first positive case of COVID-19 on the 2^nd^ of March 2020, which lead to a reduction in the daily and total new cases. Consequently, less mortality was observed in Saudi Arabia, where the estimated death rate is only (0.7%) in COVID-19 positive cases, compared to other countries of about (7%),(31). It is essential to estimate the peak-time, final size and possible new cases of the epidemic to allow the health authority to take the necessary precautions, prepare the infrastructure and provide materials needed for patients in time, to cope with the different phases of the epidemic. Furthermore, in its fight to COVID-19, the Saudi Ministry of Health introduced community testing surveillance of active screening which commenced mid-April, to discover asymptomatic cases and to improve contact tracing. Initially, they conducted 5000-6000 tests daily for a month, which eventually tripled to reach 16,000-18,000, and as of 19^th^ May, the total number of PCR-tests conducted reached 618,084 tests (32). Currently, as of 21^st^ June 2020 more than one million individuals have been screened. Early detection of the cases was the central goal of the screening; initially, over 150 teams from the Ministry of Health conveyed the visits to crowded neighborhoods and residential complexes of workers in Makkah and Madinah. The surveillance also covers Dammam, Jeddah, and Hofuf. Initially, community active screening was conducted selectively in crowded high-risk areas that reported many COVID-19 positive cases and residential buildings with suspected cases. 82% of the cases identified through active screening were clinically asymptomatic (33). Collectively, these mediations limited the spread of Covid-19 in Saudi Arabia, and eventually, the total recoveries have surpassed the number of new cases as of the last week of May 2020. Meanwhile, healthcare infrastructure was prepared by increasing ICU units and other patient’s needs to handle the epidemic waves and surges properly. Komies et al. (6) reported that the extreme 22 intervention measures imposed early by the Saudi government, led to a significant decline in the numbers of infected and mortality cases per day in Saudi Arabia compared to the UK. Subsequently, social disruption was alleviated in steps, and the lockdown of the country was eased after two months and a half (34). The reported cases reached the first observed peak on 15^th^ May then declined on the subsequent week which corresponds to the last week of Ramadan where partial curfew was implemented. Nonetheless, a second complete lockdown was imposed on the following week (corresponding to Eid Elfitr holiday) which preceded an incomplete lockdown and relaxation of the curfew which resulted in the sighted jump in the incidence of the new cases from 29^th^ May. Almost, more than three months has passed since the first reported incidence of COVID-19 in the KSA and the trend of new incidence over the last two months of April and May were observed to fluctuate remarkably with sharp spike. This could be explained in part by the introduction of active screening surveys, improvement in contact tracing and interruption of NPIs matched with intermittent episodes of relaxations. Many surges occurred during these episodes of relaxation as witnessed recently in big cities (Riyadh, Jeddah and Makkah), which led to revert to complete lockdown of Jeddah on 6^th^ June as excessive occupancy rates of ICUs were observed (35). Cultures and environment play a significant role in epidemics’ behavior, and people tend to gather on religious and social ceremonies, which might affect the prevalence of the disease. Thus, transmission of the virus increased when people didn’t abide by the rules of safety imposed by health authority guidelines specially as active screening revealed that most of the COVID-19 were asymptomatic (36). It was reported that the increase in the COVID-19 new cases observed during the last period was attributed to crowded social gatherings violating health advice and universal rules as reflected in the increase in percentages of women and children of COVID-19 positive cases. Some of these cases were linked to gathering of 30 or more people, as reveled by epidemiological investigations, which violated the precautionary rules set by KSA authorities (37). Mathematical models have always helped in short- and long-term predictions of cases and have assisted the decision-makers in their planning for the needed resources and monitoring measurements desired to control the outbreak (7, 8). ARIMA and logistic growth models have been developed to model the short and long-term forecasts, respectively of COVID-19 in KSA. Recently, ARIMA model was applied to give estimates till May 21, 2020 of daily and cumulative COVID-19 cases in KSA. But recovery and death were not forecasted in that report, as the pandemic was in its very primary phase, and the data used was till April 21,2020 (Alzahrani SI, 2020). In our study, ARIMA model was utilised for the first time to precisely estimate the short-term prevalence of recovered and deceased COVID-19 cases in KSA. The analysis, estimated that recovery and death rates to be 70.3% and 0.87% out of confirmed cases, by first of July 2020. Prevalence of daily confirmed COVID-19 cases were also estimated and predictions were given upcoming 10 days till the first of July 2020. Indeed, after 4 months of the pandemic, our models gave more accurate projections compared to the previous report (Alzahrani SI, 2020) as enough data have been accumulated which enabled adequate training of the model. ARIMA model is a linear model and has many advantages over other models in short-term forecasting but, has shortcomings when predicting longer terms events. Especially, for the trend of COVID-19 outbreak which tended to be exponential at early phases and was directly affected by the precautionary interventions imposed by the government and the compliance of the citizens. Consequently, Logistic Growth models have been developed in this study to give more realistic long-term forecast of COVID-19 pandemic in KSA. In order to deal with the fluctuating trend of COVID-19 incidence in the last three months, two different scenarios were depicted to predict the epidemic dynamics by mathematical models. The first scenario covered the period between 2^nd^ March and 28^th^ May when the first peak was observed around mid-May and dropped afterwards. Our analysis projected that the COVID-19 epidemic to end on the 4^th^ August if folks followed the recommended personal and social safety guidelines upon relaxation of curfew. The second scenario was simulated as a result of the sharp spike witnessed in the trend of the new cases on the last week of May and continue to escalate till the time of current writing-21^st^ June and estimated the epidemic to end on 29^th^ September, assuming that the NPIs will be maintained while normal life is resumed carefully. Previous reports employed Exponential Logistic and SIR models to estimating the COVID-19 dynamics in KSA in the very early phase of the epidemic before the introduction of active mass screening (6). They utilized data until the first of April 2020, and their forecasts of the epidemic’s final-size was found to be only 2064 and the pattern of recovery and mortality rates in Saudi Arabia could not be estimated, due to incomplete data at the time of their analysis (6). The predictions given by logistic growth in our study were more realistic as it employed historical data covering longer periods with more interventions, than the one used in the previous study (6). The results presented here in this report projected the epidemiological aspects of high concerns to health authority and the general public. It estimated trajectory trends and prevalence of the pandemic of COVIDs in the near future. Indeed, the models developed in this study could be used in the future for other infectious diseases epidemics. COVID-19 is caused by a novel virus with many unknown factors, and its epidemiology is associated with dynamic features. Mathematical Models are data driven projections, sensitive to alteration in the prevalence trend that may occur due to new mediations. Currently, vaccine development is in progress so, launch of a vaccine will change the scenarios, hopefully for the best, which mandate updating of the models to reflect the projection at that time.

## 4. Conclusions

Two models (ARIMA and logistic growth) were employed to predict the COVID-19 prevalence in Saudi Arabia. The analysis provided estimates of the prevalence, total size, and peak-time covering two phases COVID-19 outbreak in Saudi Arabia. If folks abide by authority precautionary recommendations and universal hygiene rules, the epidemic would have ended in 5^th^ August as predicted in the first scenario. Unfortunately, upon relaxation of the curfew during the month of May 2020, publics did not adhere to precautionary rules set by the Saudi authority. Especially, when people ignored safety guidelines upon gatherings in the holy month of Ramadan. Consequently, escalation in the number of daily confirmed cases was observed. Finally, we hope these forecasts will help the health authority and decision-makers, in their planning to allocate the resources needed to cope with the epidemics final size. Indeed, NPIs alone are not enough to manage the epidemic as it is hard to be maintained for a long time. Thus, active case screening, contact tracing and primary health education through social media campaign and local press should be enforced concomitantly with a vigilant lessening of the lockdown to prevent waves and spikes of new cases. Models should be updated in the future, with new data as the pandemic evolve to give relevant predictions specially if the vaccine become available. Further, mathematical modeling should be developed to provide short- and long-term estimates of epidemic prevalence and trend of other disease epidemics.

## Data Availability

COVID-19 data was taken from the Saudi Ministry of Health (MOH) website (Sehhtty).

https://sehhtty.com./sa-covid

## Acknowledgements

The help and support of the Research Center Administration and Research Advisory Council is highly appreciated. Specially Dr. Ali AL Zahrani Executive Director of Research Center, as he initiated a call for research on COVID-19 soon after the report of the firs case in March 2020 in KSA. Furthermore, he initiated a Fast Track Committee to facilitate the review and approval of proposals submitted to ORA. Special thanks for Dr. Ayodele Alaiya, Prof. Norma Alias and Mr. Hassan Sirelkhatim (data scientist), Mrs.Yusra Elkamali (Statistician) for critically reviewing the manuscript. Many thank Dr. Sara Elsheikh for fruitful discussion.

## Ethical Consideration

This study was conducted in compliance with the Office of Research Affairs (IRB), King Faisal Specialist and Research Center. Only free accessible data sources were used, so no ethical consideration. The study conducted is part of RAC# 2200019.

